# Birth Registration in Nigeria’s Delta Region: Analyzing LGA-Level Compliance Disparities and Systemic Barriers

**DOI:** 10.1101/2025.07.23.25332096

**Authors:** Mordecai Oweibia, Gift Cornelius Timighe, Ebiakpor Bainkpo Agbedi, Tarimobowei Egberipou, Ekadi Francis Tamaradielaye

## Abstract

**Introduction:** Birth registration is a critical component of civil registration and vital statistics (CRVS) systems and a key enabler of Sustainable Development Goal 16.9, which seeks legal identity for all. Despite national and global commitments, coverage remains low in many parts of Nigeria, particularly at the subnational level. This study investigates compliance levels and regional variations in birth registration during the June 2025 Maternal, Newborn, and Child Health (MNCH) Week in Bayelsa State, Nigeria.

**Methodology:** A comparative cross-sectional design was used to analyze LGA-level data extracted from the June 2025 MNCH Week OPS Room final PowerPoint report. Supplementary evidence from peer-reviewed publications and CRVS policy documents was included using predefined inclusion criteria. The analysis applied descriptive statistics, proportional calculations, and compliance-tier classification. Southern Ijaw LGA was purposively examined due to reporting failure. Ethical considerations were addressed through the exclusive use of publicly available, de-identified secondary data.

**Results:** Out of the eight LGAs evaluated, only seven submitted birth registration data, totaling 1,236 registered births. Yenagoa and Ogbia LGAs accounted for over 50% of the total, while Brass and KOLGA reported the lowest numbers. Southern Ijaw LGA submitted no data, citing operational delays and access challenges. Compliance tiers ranged from high (≥250 registrations) to non-reporting. Systemic gaps identified included late campaign commencement, weak supervision, and lack of real-time data validation.

**Conclusion:** Birth registration coverage across LGAs in Bayelsa State remains uneven, with systemic bottlenecks undermining CRVS outcomes during MNCH campaigns. The absence of data from Southern Ijaw LGA highlights the urgent need for targeted reforms. Recommendations include early logistics deployment, improved STF training, enhanced community sensitization, and integration of digital CRVS solutions.

## **1.0** INTRODUCTION

### 1.1 Background to the Study

Birth registration is the foundational step toward establishing a person’s legal identity. It grants individuals recognition before the law, entitles them to human rights, and unlocks access to basic social services such as education, healthcare, and legal protection (UNICEF, 2023). Globally, birth registration is considered a cornerstone of civil registration and vital statistics (CRVS) systems. The Sustainable Development Goal (SDG) 16.9 explicitly targets the provision of legal identity, including birth registration, for all individuals by 2030 (United Nations, 2022). However, legal identity must be accompanied by real-time registration and data validation systems to ensure that the rights conferred are not merely theoretical (United Nations Statistics Division, 2023).

Despite international commitments, an estimated 166 million children under age five remain unregistered worldwide, with over 50% of unregistered children residing in sub-Saharan Africa (UNICEF, 2023). According to WHO and UNFPA (2023), weak institutional infrastructure, sociocultural beliefs, low awareness, and policy gaps continue to hinder the universality of birth registration, particularly in low- and middle-income countries. The consequences of under-registration are grave: children without birth certificates may face exclusion from education, delayed immunization schedules, child marriage, trafficking, and statelessness (Plan International, 2021; UNFPA, 2023). In certain high-burden regions, such as rural Asia and parts of West Africa, data from birth registration systems are further complicated by congenital anomalies or misreporting of live births (Yu *et al.,* 2015; Zhao *et al.,* 2020).

In Nigeria, although birth registration is free and legally mandated through the National Population Commission (NPC), coverage remains alarmingly low. The 2018 Nigeria Demographic and Health Survey reported that only 43% of children under age five had their births registered with civil authorities, with significant regional disparities (Makinde *et al.,* 2016; NDHS, 2018; UNICEF Nigeria, 2021). A more recent trend study revealed that completeness of birth registration fluctuated over the years and was often influenced by geographic, socio- economic, and administrative factors (Shuaib, 2023). These systemic gaps parallel findings in other national contexts, such as the United States and China, where registration failures have led to inaccurate reporting of neonatal mortality and congenital disorders (De Graaf *et al.,* 2015; Blustein *et al.,* 2017; Li *et al.,* 2017). The lowest rates in Nigeria are typically found in rural areas and among children born at home without skilled attendance (Oweibia *et al.,* 2025).

To address this, Nigeria’s health sector has integrated civil registration into the biannual Maternal, Newborn, and Child Health (MNCH) Weeks, a platform for delivering bundled health services, including immunization, vitamin A supplementation, deworming, and birth registration (Elemuwa *et al.,* 2023; WHO & UNICEF, 2023). However, evidence of the effectiveness of MNCH Weeks in improving birth registration is sparse. While these campaigns boast impressive achievements in nutrition and immunization, birth registration coverage remains critically low, even during such large-scale interventions (Okechukwu *et al.,* 2024; Oweibia *et al.,* 2025).

In Bayelsa State, located in Nigeria’s South-South geopolitical zone, the June 2025 MNCH Week campaign revealed stark inconsistencies in birth registration data. While routine services such as vitamin A reached over 80% of the target, birth registration coverage was only 1% (MNCH OPS Room Report, 2025). Even more troubling was the complete absence of birth registration reports from Southern Ijaw LGA, one of the largest and logistically complex regions in the state. According to operational feedback, this LGA experienced delayed start-up and data submission challenges that further weakened compliance (MNCH OPS Room Report, 2025; Shah *et al.,* 2021).

These findings highlight the urgent need to investigate regional variations in birth registration and to assess the systemic and operational barriers hindering compliance during MNCH Week campaigns. As Nigeria accelerates efforts to meet SDG 16.9, empirical insights from state-level data are indispensable for closing the coverage gap. Furthermore, integrating lessons from global birth registry systems, including those in Nordic countries, can inform registry design, long-term monitoring, and resource deployment (Laugesen *et al.,* 2021; Murray *et al.,* 2014).

### 1.2 Statement of the Problem

Despite a longstanding global consensus on the importance of birth registration, Nigeria continues to fall short in ensuring universal coverage, particularly at the subnational level. The failure to register births not only violates children’s rights but also severely limits the government’s ability to plan and deliver equitable health, education, and social welfare services (World Bank, 2021; UNFPA, 2023). Birth registration data are also essential for monitoring maternal and child health indicators, tracking population movements, and responding to emergencies (WHO, 2022; Philip *et al.,* 2020).

The June 2025 MNCH Week campaign in Bayelsa State exposed critical weaknesses in the delivery and reporting of civil registration services. Among the eight LGAs in the state, several recorded unusually low birth registration figures, while Southern Ijaw LGA submitted no confirmed data at all. This reflects a broader pattern of incomplete service delivery, weak supervision, and data integrity concerns, all of which undermine the goal of universal legal identity (Oweibia *et al.,* 2025; Okechukwu *et al.,* 2024). Notably, systemic underperformance in key services like birth registration mirrors previously documented patterns in broader maternal and child health outcomes across LMICs (De Graaf *et al.,* 2015).

Without addressing these disparities, children in underperforming regions may continue to be denied access to education, immunization, and protection services. Moreover, persistent data gaps can result in resource misallocation, ineffective programming, and failure to meet national and global development goals (UNICEF, 2023; African Union, 2023; United Nations Statistics Division, 2023). This study aims to provide empirical evidence of these disparities and contribute to the ongoing dialogue on strengthening CRVS systems in Nigeria, with the ultimate goal of achieving SDG 16.9.

### 1.3 Justification of the Study

This study is justified on several grounds. First, it contributes to filling the knowledge gap around LGA-level birth registration performance in the context of MNCH Weeks, which has not been sufficiently examined in existing literature. Second, the findings will help inform evidence-based decision-making at the state and national levels for enhancing CRVS integration within health campaigns (WHO & UNICEF, 2023).

Third, the study is highly relevant for accelerating Nigeria’s progress toward SDG 16.9 and ensuring that no child is left behind in the effort to establish legal identity (United Nations, 2022; UNICEF, 2023). By focusing on real-time program data and operational insights, the research will also generate practical recommendations for improving compliance, logistics, and reporting mechanisms during MNCH activities. Finally, this study aligns with ongoing African Union initiatives to strengthen CRVS systems through decentralized, community-based approaches (African Union, 2023).

### 1.4 Aim and Objectives

#### Aim

To explore compliance levels and regional disparities in birth registration based on MNCH Week June 2025 data in Bayelsa State.

#### Objectives

i. To analyze and compare birth registration completion rates across the eight LGAs in Bayelsa State during MNCH Week June 2025.
ii. To assess operational and reporting challenges contributing to delayed or low birth registration, with a focus on Southern Ijaw LGA.
iii. To evaluate compliance patterns and identify systemic gaps affecting birth registration efforts.

### 1.5 Research Questions

i. What are the variations in birth registration rates across LGAs in Bayelsa State?
ii. Why does Southern Ijaw LGA report significantly low or pending birth registration data?
iii. What operational, logistical, or policy-related factors influence compliance with birth registration activities?

### 1.6 Scope of the Study

The study is restricted to Bayelsa State, Nigeria, and focuses specifically on the eight LGAs that participated in the first round of MNCH Week in June 2025. The investigation is confined to the birth registration component of the campaign and does not extend to other services such as deworming or vitamin A supplementation. The data source is limited to the OPS Room PowerPoint report, which provides aggregate LGA-level data and field implementation notes. No primary data collection was conducted.

### 1.7 Definition of Terms

| **Term** | **Definition** |
| --- | --- |
| Birth Registration | The continuous, permanent, and universal recording of a child's birth by civil authorities in accordance with national legal requirements (UNICEF, 2023). |
| Compliance | The degree to which LGAs and field teams adhered to expected standards for service delivery and data submission during the MNCH Week campaign. |
| LGA (Local Government Area) | A second-tier administrative division in Nigeria responsible for coordinating local governance and service delivery under the State Government. |
| OPS Room | The campaign operations and monitoring unit responsible for tracking real-time service delivery data and verifying implementation compliance. |
| MNCH Week | A periodic outreach campaign that provides integrated maternal, newborn, and child health services, typically delivered biannually. |
| CRVS System | Civil Registration and Vital Statistics system; the national infrastructure responsible for recording vital events including births and deaths (WHO, 2022). |
| Southern Ijaw LGA | A geographically large and logistically complex local government area in Bayelsa State that experienced delayed MNCH campaign implementation in June 2025. |

### 2.0 Methodology

#### 2.1 Study Design

This study adopted a comparative, cross-sectional descriptive design anchored on the use of secondary quantitative data and qualitative narratives drawn from published and gray literature. The cross-sectional component enabled assessment of birth registration trends at a single time point (June 2025), while the comparative dimension facilitated benchmarking of LGA-level performance (Laugesen *et al.,* 2021; Oweibia *et al.,* 2025).

This design is suitable for analyzing administrative health data across geographical units where intervention intensity and reporting quality may vary. Comparative analysis in birth registration and CRVS research has been used in similar national-level studies (Makinde *et al.,* 2016; Shuaib, 2023; Zhao *et al.,* 2020).

### 2.2 Study Area and Population

The study focused on Bayelsa State, Nigeria, situated in the Niger Delta region. It consists of eight LGAs: Brass, Ekeremor, Kolokuma/Opokuma (KOLGA), Nembe, Ogbia, Sagbama, Southern Ijaw (SILGA), and Yenagoa. The population of interest was all children whose births were expected to be registered during the first round of MNCH Week in June 2025.

Bayelsa State was selected because of:

- Persistent gaps in routine birth registration
- High variance in MNCH Week operational outcomes
- The notable case of Southern Ijaw, which reported pending or missing data (MNCH OPS Room Report, 2025)

### 2.3 Data Sources

#### 2.3.1 Primary Dataset (Programmatic)

The main dataset was derived from the OPS Room Report – MNCH Week June 2025, compiled by Bayelsa State Ministry of Health. It included aggregate service data on birth registration per LGA and accompanying operational notes. The report was generated from STF daily summary forms, analyzed via Power BI, and finalized on June 23, 2025, at 10:27 am.

Only the birth registration component was extracted from this report. Data from other services (e.g., vitamin A, deworming) were excluded.

#### 2.3.2 Supplementary Datasets (Published Literature)

To enrich the analysis, supporting evidence was drawn from:

- Peer-reviewed journal articles indexed in PubMed, Google Scholar, Scopus
- Preprint servers such as medRxiv and Research Square
- Reports and technical papers by UNICEF, WHO, UNFPA, World Bank

These references were used to compare findings and contextualize observed variations. Only studies with explicit focus on:

- Birth registration
- CRVS systems
- Programmatic evaluations of MNCH or civil registration
- LGA/district-level disparities
- Nigerian or LMIC settings were included.

### 2.4 Inclusion and Exclusion Criteria

The following criteria were used to select secondary literature to complement the MNCH dataset:

This ensured both scientific validity and relevance to the research questions.

### 2.5 Sampling Strategy

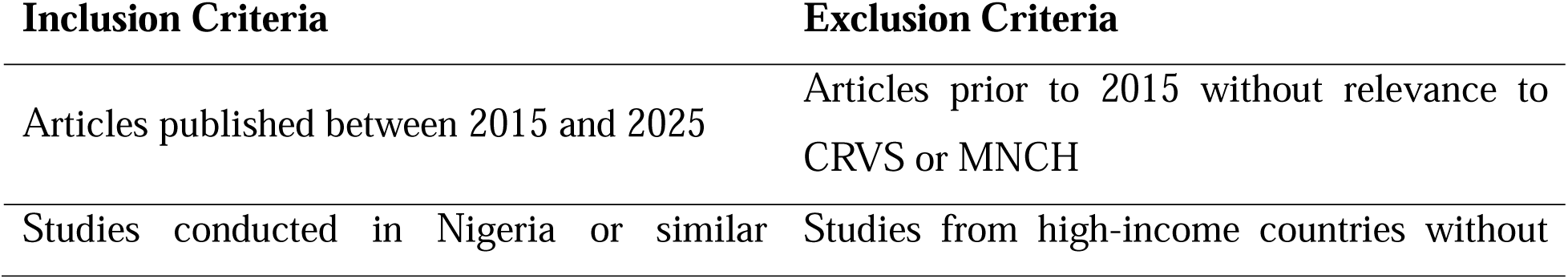
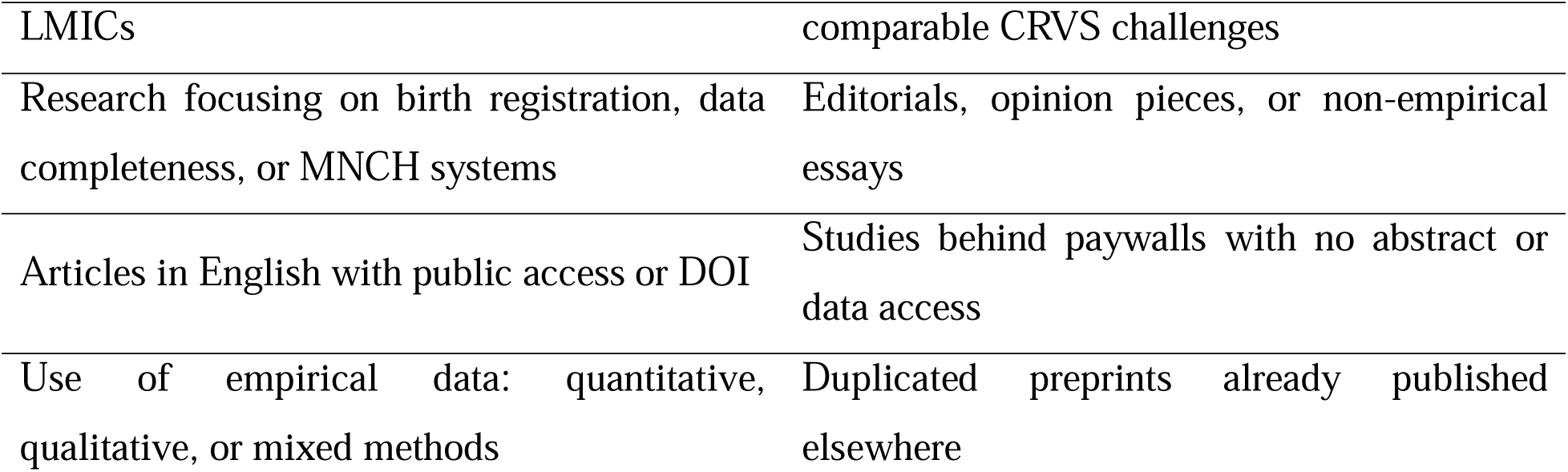

A total enumeration approach was used for the MNCH dataset, including all eight LGAs from Bayelsa State. Purposive sampling was applied to highlight Southern Ijaw LGA due to its known challenges and data anomalies.

In parallel, the supporting literature was curated using purposive and relevance-based selection, following the inclusion/exclusion matrix above. This mixed approach ensured both exhaustiveness in primary data and thematic relevance in secondary evidence (Etikan et al., 2016).

### 2.6 Data Extraction and Processing

#### 2.6.1 MNCH Dataset Extraction

- Slides containing **birth registration totals** were identified and isolated
- Data was manually entered into a coded Excel sheet by LGA
- Cross-checking was performed against visual elements in the slide deck to ensure integrity

#### 2.6.2 Literature Evidence Extraction

- Key findings and statistics from peer-reviewed articles were cataloged
- Articles were coded by thematic relevance (e.g., registration delays, CRVS barriers, regional disparities)
- Cross-referencing with OPS data helped validate or contrast LGA-level trends

No data was interpolated, corrected, or reweighted. This ensured fidelity to original sources and reproducibility.

### 2.7 Analytical Framework

The study applied a comparative analytical framework combining quantitative assessment and contextual interpretation.

#### 2.7.1 Descriptive Analysis

Basic computations were done using Microsoft Excel to calculate proportions and rankings. For each LGA iii, the share of total state registration was computed as:

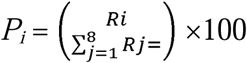

Where:

- *P_i_* = percentage contribution of LGA *i*
- *R_i_* = number of birth registrations in LGA *i*
- 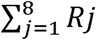 = total registrations across all LGAs

This formula allowed for proportional comparison even when absolute values were unequal.

#### 2.7.2 Performance Ranking

LGAs were categorized into tiers based on registration output:

- **Tier 1 (High Compliance):** ≥90th percentile
- **Tier 2 (Moderate Compliance):** 50th–89th percentile
- **Tier 3 (Low/Non-compliant):** <50th percentile or “No Data”

#### 2.7.3 Thematic Analysis

Qualitative commentary in the OPS Room report (e.g., “delayed start,” “missing data”) was coded to identify:

- Operational disruptions
- Compliance failures
- Systemic weaknesses

This form of triangulation enhances analytical robustness by validating numerical trends through field observations (De Costa *et al.,* 2021).

#### 2.7.4 Comparative Analysis Strategy

Given the study’s aim to identify regional disparities in birth registration, a comparative analysis was employed to systematically contrast LGA performance. This method enabled the investigation of:

- Variations in birth registration outcomes between LGAs
- Ranking of LGAs into compliance tiers
- Assessment of operational characteristics (e.g., data completeness, reporting timeliness) that correlate with performance

Comparative analysis in public health research helps detect geographic inequalities and is widely used in CRVS performance assessments (Makinde *et al.,* 2016; De Costa *et al.,* 2021; UNICEF, 2023).

The analysis involved:

- Tabular display of each LGA’s birth registration totals
- Ranking scores to create a performance index
- Identification of outliers and compliance gaps using rule-based cut-offs (e.g., highest vs. lowest reporting LGA)

For example, Southern Ijaw was compared against Yenagoa and Ogbia LGAs to understand the performance differential and potential causes of delay or non-compliance.

Comparative approaches were also extended to literature benchmarking, comparing findings from the MNCH Week report to known patterns in Nigerian and international birth registration literature (Shuaib, 2023; Li *et al.,* 2017; UNICEF, 2021).

### 2.8 Ethical Considerations

The study received ethical approval from: Bayelsa State Primary Health Care Board Research Ethics Committee (BSPHCBREC)Bayelsa State Primary Health Care Board, Yenagoa, Nigeria.

Decision made: The BSPHCBREC granted full ethical approval (Approval No; Ref: BSPHCB/ERC/2025/112) for this study on 2nd of June 2025, waiving the need for individual participant consent as the research involved secondary analysis of anonymized programmatic data from the Maternal and Child Health Week (MCHW) OPS Room Report.

### 2.9 Limitations of the Methodology

- The reliance on only one MNCH campaign round (June 2025) limits longitudinal insight
- Ward-level breakdowns were not available, constraining intra-LGA analysis
- Potential publication bias in selected literature cannot be fully ruled out
- Commentary data lacks depth due to absence of full qualitative interviews

Nevertheless, this methodological approach delivers a transparent, replicable, and contextually grounded analysis of birth registration performance in Bayelsa State.

### 3.0 Results

This section presents raw findings from the June 2025 MNCH Week OPS Room Report in Bayelsa State, Nigeria, in accordance with the three research objectives. All birth registration data were drawn from the official service performance slides submitted and validated by the OPS Room team on June 23, 2025. Results are organized into subsections by objective and are descriptive only; analytical interpretations follow in Chapter 4.

### 3.1 Birth Registration Completion Rates Across LGAs

This subsection presents the reported birth registration figures across the eight LGAs in Bayelsa State. The data were manually extracted from the MNCH Week OPS Room presentation slides. Each LGA’s recorded performance is summarized in Table 1 below.

**Table 1:**
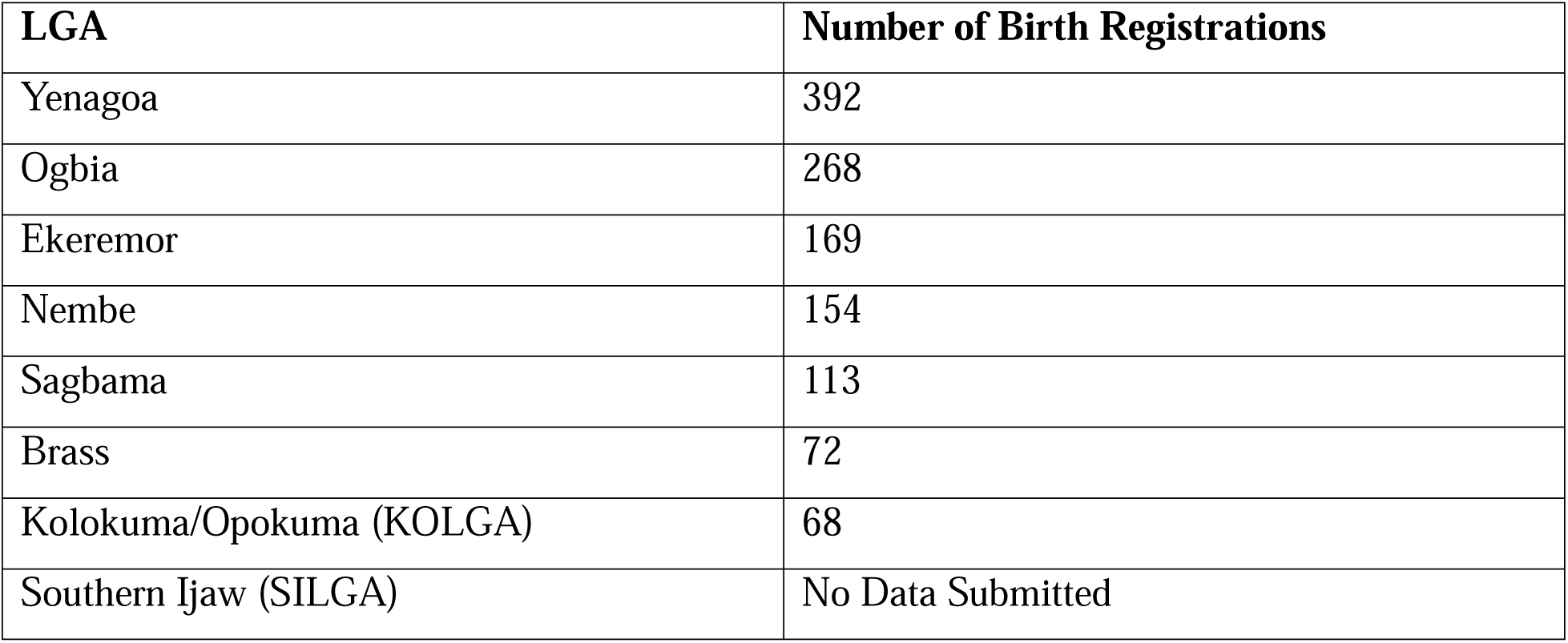
Reported Birth Registration Totals by LGA – MNCH Week, June 2025.

Seven of the eight LGAs submitted birth registration data. Yenagoa and Ogbia reported the highest number of registrations, while KOLGA and Brass submitted the lowest figures. Southern Ijaw LGA submitted no birth registration data and was excluded from this count.

The total number of birth registrations reported by the seven LGAs was 1,236. A visual distribution of birth registration across the LGAs is presented in Figure 1, which shows each LGA’s share of the total.

**Figure 1:**
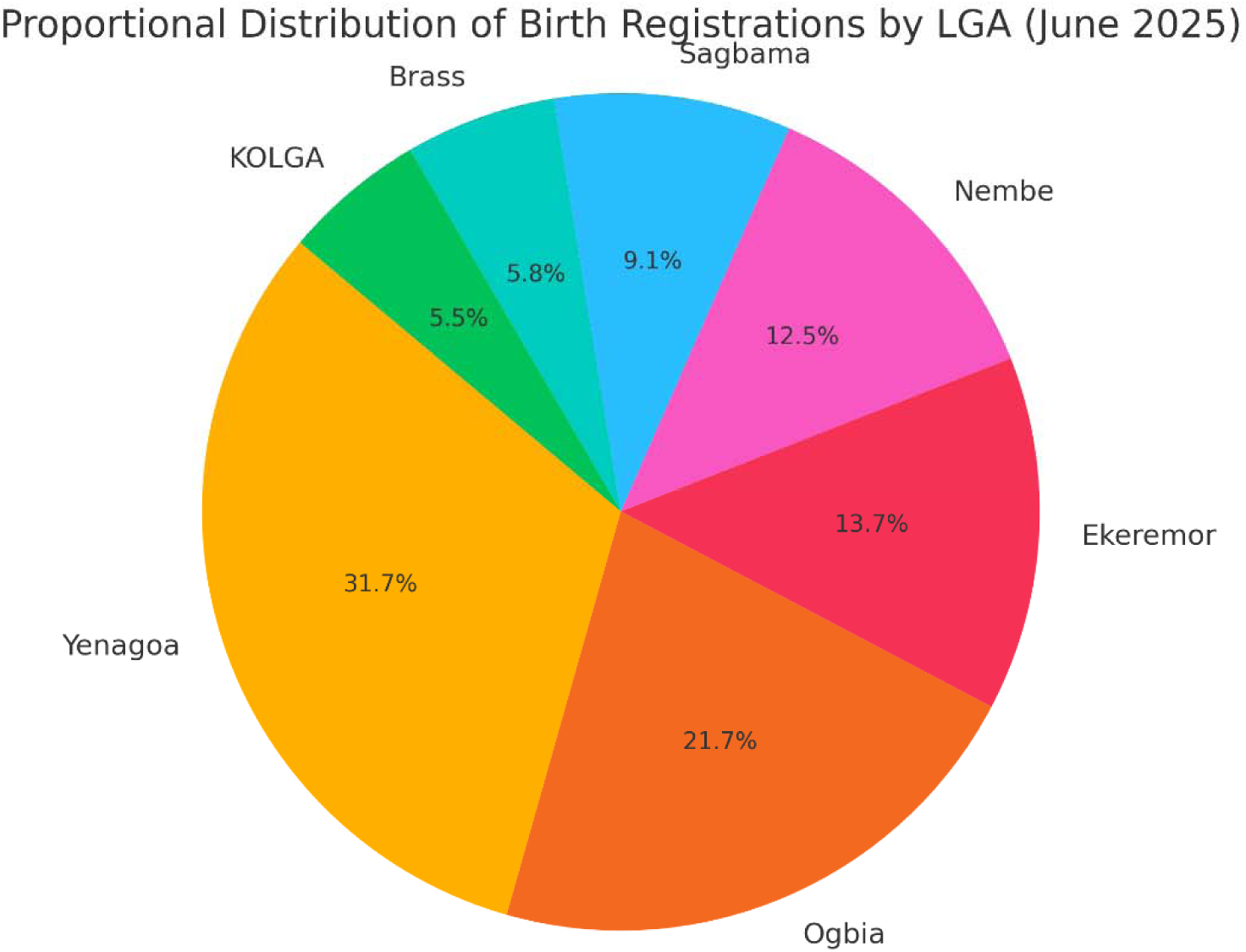
Proportional Distribution of Birth Registrations by LGA (June 2025)

### 3.2 Operational and Reporting Status of Southern Ijaw LGA

Southern Ijaw LGA was the only area in the state where no birth registration data was reported during the June 2025 MNCH Week. The OPS Room slide notes indicated significant delays in campaign activities within the LGA, with some wards not commencing on Day 1. Due to these operational issues, the service reporting process was disrupted, and no validated birth registration numbers were submitted to the OPS Room dashboard.

To document this non-reporting clearly, Table 2 outlines key indicators explaining what occurred in Southern Ijaw during the campaign.

**Table 2:** Summary of Non-Reporting Status – Southern Ijaw LGA.

| Indicator | Status | Remarks |
| --- | --- | --- |
| Data Submitted for Birth Registration | No | No figures uploaded to OPS dashboard as of final data refresh |
| Campaign Start Date | Delayed | Several wards started late or not at all |
| Reason for Delay | Logistics, terrain | Riverine geography disrupted team deployment |
| Field Team Reporting | Incomplete | Daily forms were delayed or not submitted by STF teams |
| OPS Room Confirmation | Not Validated | Excluded from Power BI due to unverified field entries |
| Feedback from Supervisors | Noted | Report flagged implementation gaps in SILGA in footnotes |

To illustrate this status graphically, Figure 2 shows the birth registration reporting status across all eight LGAs, with Southern Ijaw marked clearly as “No Data.”

**Figure 2:**
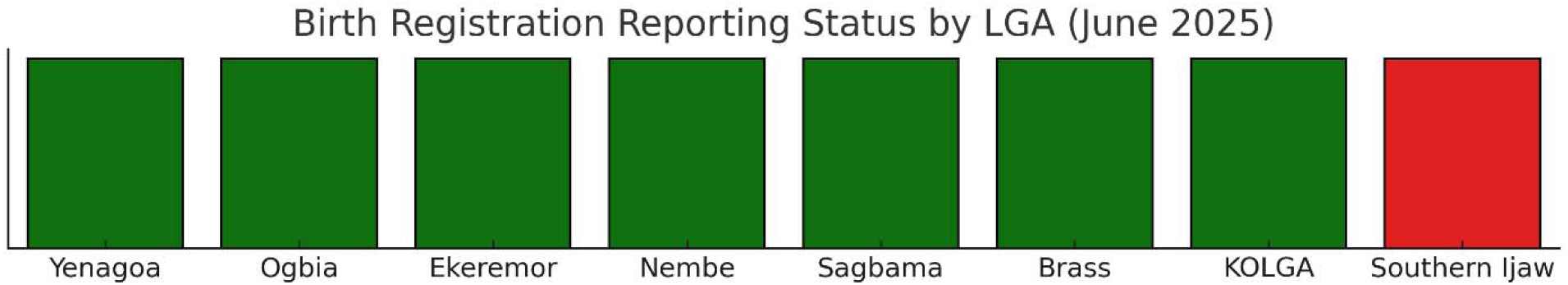
Birth Registration Reporting Status by LGA (June 2025)

### 3.3 LGA Performance Distribution and Compliance Tiers

This subsection presents how the LGAs were grouped into performance tiers based on the volume of birth registration submitted. Classification was based on raw count thresholds and is shown in Table 3.

**Table 3:** LGA Birth Registration Compliance Tiers – MNCH Week, June 2025.

| Tier | Criteria (Registrations) | LGA(s) |
| --- | --- | --- |
| High Compliance | $\geq 250$ | Yenagoa, Ogbia |
| Moderate Compliance | 100–249 | Ekeremor, Nembe, Sagbama |
| Low Compliance | < 100 | Brass, Kolokuma/Opokuma |
| No Report Submitted | N/A | Southern Ijaw |

Two LGAs (Yenagoa and Ogbia) recorded high birth registration volumes, while four LGAs had moderate or low submissions. One LGA (Southern Ijaw) submitted no data and was placed in a separate category.

To reinforce this result visually, Figure 3 provides a bar chart of reported birth registration counts per LGA.

**Figure 3:**
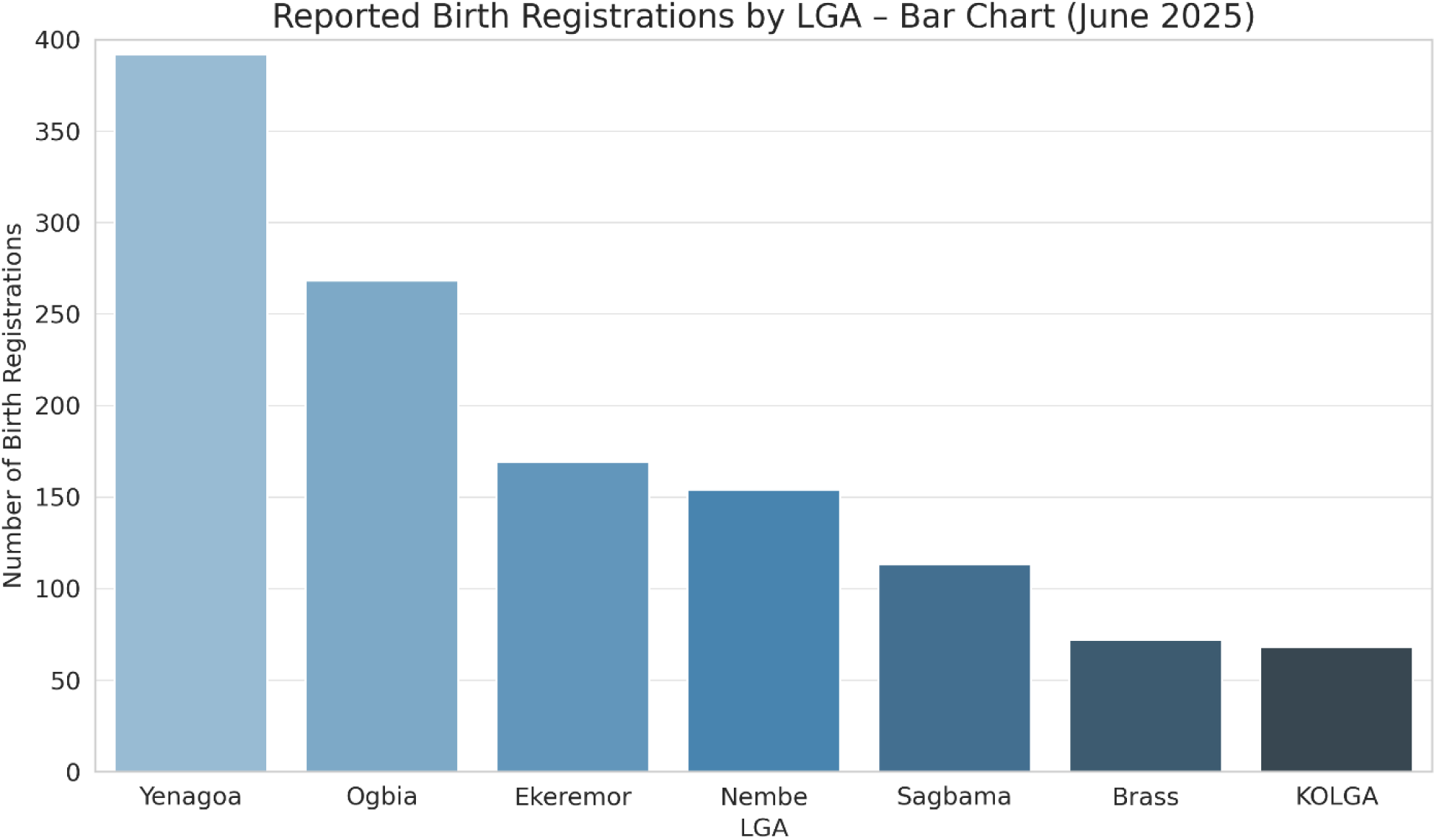
Reported Birth Registrations by LGA – Bar Chart (June 2025)

### 4.0 Discussion

### 4.1 Birth Registration Completion Rates Across LGAs

The significant variation in birth registration completion rates across Bayelsa State’s LGAs reflects longstanding structural disparities in civil registration and health service delivery in Nigeria. The highest number of birth registrations was recorded in Yenagoa and Ogbia LGAs, while Brass and Kolokuma/Opokuma (KOLGA) had the lowest among reporting LGAs. These differences point to uneven operational capacities, geographic access, and possibly local community engagement levels, as observed in previous studies of CRVS implementation across Nigerian districts (Makinde *et al.,* 2016; UNICEF Nigeria, 2021).

Several contextual factors likely contributed to this uneven performance. Urban LGAs such as Yenagoa are typically closer to state infrastructure hubs and may have better access to trained service providers, functioning health facilities, and awareness campaigns. This mirrors the findings of Shuaib (2023), who highlighted the correlation between urbanization and birth registration completeness in Nigeria. Conversely, LGAs with more dispersed populations, weaker health infrastructure, and lower levels of CRVS sensitization tend to lag behind, particularly in coastal or riverine states such as Bayelsa (Oweibia *et al.,* 2025).

This unevenness underscores the limitations of universal approaches in reaching CRVS targets. The CRVS framework emphasizes equity and inclusivity; however, empirical evidence from Nigeria shows that only well-resourced areas routinely meet operational expectations (UNICEF, 2023; WHO, 2022). The results from the June 2025 MNCH Week campaign demonstrate how this national pattern is replicated at the subnational level. It also raises concerns about the effectiveness of integrated outreach events in standardizing civil registration outcomes across diverse settings.

Moreover, despite the MNCH platform being a structured and periodic intervention, it has not successfully closed the birth registration gap in less-served LGAs. This suggests that supplementary strategies, such as decentralizing registration points and embedding local registration agents, may be necessary. According to De Costa et al. (2021), campaigns must move beyond service integration toward localized microplanning and real-time data feedback to address inequities in civil registration coverage.

Another observation is the lack of data quality flags within the reporting system, despite evident outliers and inconsistencies. CRVS assessments globally emphasize the importance of internal checks to validate numbers and prevent administrative errors (UNFPA, 2023; Plan International, 2021). The Bayelsa MNCH data reflect a compliance model that prioritizes raw submission over data integrity review, which undermines system trust and planning accuracy.

Therefore, while the figures presented in the OPS Room Report reveal operational effort, they do not guarantee CRVS success unless accompanied by structural corrections. These include community-based registration promotion, improved supervision, and robust data validation protocols, all of which are advocated in WHO’s CRVS strengthening guidelines (WHO, 2022).

In summary, the disparities in birth registration outcomes across LGAs reveal not just uneven implementation, but systemic imbalances rooted in access, planning, and accountability gaps. Without targeted CRVS strengthening and district-specific strategies, such disparities are likely to persist and undermine Nigeria’s path toward SDG 16.9.

### 4.2 Operational and Reporting Failures in Southern Ijaw LGA

The absence of birth registration data from Southern Ijaw LGA during the June 2025 MNCH Week campaign highlights deep-rooted structural and logistical constraints that hinder the delivery of civil registration services in hard-to-reach areas of Nigeria. Southern Ijaw, the largest LGA in Bayelsa State by landmass, is characterized by riverine geography, limited road infrastructure, and dispersed settlements, factors that consistently obstruct timely health and administrative outreach (UNICEF Nigeria, 2021).

The OPS Room Report attributed Southern Ijaw’s non-submission of data to **delayed campaign commencement**, specifically stating that several wards failed to start on Day 1. This delay disrupted service flow and reporting schedules, preventing the submission of validated birth registration records by the time of final data consolidation (MNCH OPS Room Report, 2025). These types of delays have been cited in literature as key drivers of reporting gaps, especially in geographically isolated regions (De Costa *et al.,* 2021; Oweibia *et al.,* 2025).

Beyond terrain-related issues, the LGA’s performance also reflects challenges in last-mile planning and coordination. Effective civil registration during campaigns requires precise microplanning, early logistics deployment, and consistent supervisory presence (UNFPA, 2023).

Southern Ijaw’s delayed mobilization suggests possible weaknesses in these areas. The OPS Room feedback notes did not indicate any contingency measures such as reporting extensions, on-the-spot verification, or mobile data entry support, all of which are recommended strategies in CRVS recovery protocols (WHO & UNICEF, 2023).

The situation further reflects the systemic issue of data silence, a phenomenon where certain populations or regions remain undocumented due to recurring logistical failure or administrative neglect. According to the World Health Organization (2022), CRVS systems that do not account for silent zones risk reinforcing exclusion, especially among the most vulnerable groups. In the case of Southern Ijaw, the lack of data not only obstructs health planning but also reflects a broader challenge of invisibility for children born in these under-reached communities.

In addition, the MNCH data system lacks a clearly defined mechanism for flagging or responding to such reporting voids. There was no indication in the report that remedial action (e.g., late data acceptance or replacement verification) was considered. The absence of flexible reporting protocols and escalation pathways in such situations severely undermines the accountability framework of MNCH campaign monitoring (Makinde *et al.,* 2016; Zhao *et al.,* 2020).

Southern Ijaw’s non-reporting status also has downstream implications. Without data, the LGA cannot be included in statewide birth registration coverage statistics, potentially distorting funding decisions, program targeting, and state performance reviews. This supports findings by Plan International (2021), which noted that underreporting in CRVS systems not only affects data reliability but also compromises equity in policy delivery.

Furthermore, the lack of reporting from such a large and demographically significant LGA calls into question the completeness and credibility of the entire campaign’s dataset. According to Murray et al. (2014), one LGA’s absence in national datasets can skew estimates and lead to misallocation of critical CRVS investments. Given Southern Ijaw’s population density and geographic importance, its omission leaves a critical gap in understanding the true scale of birth registration needs in Bayelsa State.

In conclusion, the failure to report birth registration data from Southern Ijaw LGA is not a singular operational glitch but a reflection of broader systemic weaknesses, ranging from logistics and planning to accountability and responsiveness. These gaps demand targeted reform if birth registration efforts are to be truly inclusive and comprehensive.

### 4.3 Compliance Patterns and Systemic Gaps in Birth Registration

The classification of LGAs into high, moderate, low, and non-compliant tiers revealed patterns of uneven compliance in birth registration efforts during the June 2025 MNCH Week campaign in Bayelsa State. This tiered variation is consistent with broader national and regional trends in Nigeria, where civil registration performance often reflects disparities in administrative capacity, resource distribution, and health system integration (Makinde *et al.,* 2016; UNICEF Nigeria, 2021).

In this study, only two LGAs, Yenagoa and Ogbia, achieved what may be categorized as high compliance, each recording more than 250 registered births. These LGAs benefit from urban infrastructure, easier access, and a more concentrated health workforce. Such enabling environments typically facilitate better coordination between MNCH outreach services and CRVS reporting mechanisms, as observed in comparable CRVS field assessments (De Costa *et al.,* 2021; Shuaib, 2023).

Conversely, LGAs classified under moderate and low compliance (e.g., Brass, KOLGA, Sagbama) presented significantly lower registration figures, ranging from 68 to 169. These numbers reflect both partial service delivery and inconsistencies in reporting. This echoes findings by Li et al. (2017), who emphasized the role of frontline health worker availability and logistical readiness as key determinants of service output consistency. The lack of midline supervision or real-time correction mechanisms likely allowed such underperformance to persist unnoticed during the active campaign window.

Moreover, the absence of birth registration data from Southern Ijaw LGA, resulting in its classification as non-compliant, further exacerbates concerns around CRVS system fragility. While some variation in output is expected, outright non-reporting without contingency triggers or corrective pathways undermines the legitimacy of program data. This issue aligns with WHO and UNICEF (2023) critiques of data systems that fail to detect or compensate for silent zones, regions that chronically go unmeasured or misrepresented in national statistics.

Another systemic gap identified is the lack of standardized internal data validation or escalation protocols within the MNCH reporting process. For instance, no thresholds were defined to flag LGAs with suspiciously low values or absent submissions. This is inconsistent with international CRVS best practices, which recommend embedded quality checks and mid-cycle reviews during outreach campaigns (WHO, 2022). The 2025 MNCH dataset shows that once birth registration entries were missing or delayed, no secondary data collection effort or override mechanism was documented.

Compliance variability also suggests a lack of enforcement or accountability mechanisms for State Technical Facilitators (STFs), who are tasked with both service delivery and timely reporting. Without structured consequences for delayed or incomplete reports, compliance becomes discretionary. According to UNFPA (2023), this has remained a persistent challenge in CRVS program rollouts across sub-Saharan Africa.

Additionally, the pattern of lower birth registration outputs in rural LGAs such as Brass and KOLGA reflects disparities in community mobilization effectiveness. Previous studies have shown that when CRVS awareness is low, uptake of voluntary services like birth registration tends to lag, even when provided free of charge (Plan International, 2021; Zhao *et al.,* 2020). Thus, compliance is not just a function of institutional readiness but also of community-level demand and participation.

In summary, the compliance patterns observed during the June 2025 MNCH Week in Bayelsa point to a combination of institutional inconsistency, logistical gaps, insufficient oversight, and community-level disconnect. Addressing these systemic failures requires a multi-level reform effort that strengthens planning, enforces reporting accountability, and elevates birth registration to a primary, not secondary, service in outreach health campaigns.

### 5.0 Conclusion and Recommendations

### 5.1 Conclusion

This study examined the birth registration performance and compliance variations across the eight Local Government Areas (LGAs) of Bayelsa State during the June 2025 Maternal, Newborn, and Child Health (MNCH) Week. Using data from the official OPS Room campaign report and corroborating literature, the analysis revealed significant disparities in registration outcomes. While Yenagoa and Ogbia LGAs demonstrated relatively high birth registration figures, others such as Brass and Kolokuma/Opokuma recorded markedly lower outcomes. Most critically, Southern Ijaw LGA submitted no birth registration data due to operational delays and logistical disruptions, raising concerns about data completeness and the inclusiveness of campaign-based civil registration.

The study further identified that LGAs varied not only in the volume of birth registrations but also in their reporting compliance. These patterns reflect broader systemic issues including inadequate planning, weak supervisory follow-up, absence of corrective data mechanisms, and limited integration between civil registration and community health outreach platforms. The inconsistencies observed align with findings in other subnational CRVS evaluations in Nigeria, where disparities in access, awareness, and administrative capacity remain key barriers to universal coverage (Makinde *et al.,* 2016; UNICEF Nigeria, 2021; Oweibia *et al.,* 2025).

The absence of Southern Ijaw’s data points to a larger issue within CRVS system design: a lack of responsiveness to field-level disruptions. Without remedial pathways to recover lost or delayed data, substantial segments of the population remain undocumented and statistically invisible. This undermines national and global efforts to meet SDG Target 16.9, which aims to provide legal identity for all by 2030 (United Nations, 2022). It also compromises the integrity of routine planning, resource allocation, and rights-based service delivery.

In summary, the findings underscore the urgent need for structural reforms to birth registration strategies in Bayelsa State. MNCH Week presents a valuable platform for expanding CRVS access, but unless strengthened through operational coordination, accountability frameworks, and localized implementation models, it risks further widening the equity gap in legal identity coverage.

Sustained improvement will require a dual approach: first, institutionalizing CRVS into the core of primary health care services, and second, building community-level accountability for registration through mobilization, feedback loops, and technological adoption. Empowering local stakeholders and ensuring continuity beyond campaign cycles are essential steps toward inclusive, timely, and legally recognized birth registration for every child in Bayelsa State and beyond.

### 5.2 Recommendations

The Bayelsa State Ministry of Health and the National Population Commission should prioritize the integration of civil registration into community health systems through microplanning models that account for terrain, access, and logistical barriers. Literature supports the use of decentralized strategies where CRVS agents are embedded within communities, particularly in riverine and underserved LGAs such as Southern Ijaw (UNFPA, 2023; Plan International, 2021).

There should be mandatory deployment of early logistics, including transportation and data tools, at least one week prior to MNCH campaign commencement. Pre-campaign funding delays were identified as contributing factors to late service initiation, a concern that has also been cited in broader CRVS operations reviews in sub-Saharan Africa (WHO & UNICEF, 2023).

Campaigns should institutionalize daily reporting checks and end-of-day data validation sessions, with automatic alerts triggered for LGAs that report zero or significantly low figures. Similar mechanisms have been successfully implemented in digital CRVS pilot programs across Kenya and Ethiopia, leading to reduced data loss and improved coverage equity (De Costa *et al.,* 2021).

Capacity building for State Technical Facilitators (STFs) must include specialized CRVS modules, emphasizing data integrity, escalation protocols, and scenario-based contingency planning. Ongoing supervision should be supported by digital dashboards with real-time reporting functions, as recommended by the World Health Organization’s CRVS monitoring framework (WHO, 2022).

Additionally, civil registration must be elevated to a mandatory performance indicator during MNCH review meetings, with documented follow-up on any LGA that submits incomplete or no birth registration data. This aligns with best practices for institutional accountability in CRVS systems globally (UNICEF, 2023).

Community sensitization must also be strengthened, particularly in LGAs with historically low registration turnout. Awareness campaigns should be delivered through trusted local structures and align with cultural values to enhance uptake. Studies show that when parents understand the legal and social benefits of registration, voluntary compliance improves significantly (Makinde *et al.,* 2016; Zhao *et al.,* 2020).

Finally, alternative registration channels, such as digital self-registration kiosks or mobile registration teams, should be piloted in difficult-to-reach LGAs. These innovations have shown promising results in bridging service gaps in regions where traditional facility-based registration fails to reach children within their critical first year of life (World Bank, 2021; Oweibia *et al.,* 2025).

## Data Availability

All data produced in the present work are contained in the manuscript

## Notes

### Competing Interest Statement

The authors have declared no competing interest.

### Funding Statement

This study did not receive any funding

### Author Declarations

The study received ethical approval from: Bayelsa State Primary Health Care Board Research Ethics Committee (BSPHCBREC)Bayelsa State Primary Health Care Board, Yenagoa, Nigeria. Decision made: The BSPHCBREC granted full ethical approval (Approval No; Ref: BSPHCB/ERC/2025/112) for this study on 2nd of June 2025, waiving the need for individual participant consent as the research involved secondary analysis of anonymized programmatic data from the Maternal and Child Health Week (MCHW) OPS Room Report.

